# Is therapeutic inertia present in primary diabetes care in Malaysia? A prospective study

**DOI:** 10.1101/19002584

**Authors:** Boon-How Chew, Husni Hussain, Ziti Akthar Supian

## Abstract

**Aims:** This prospective study aimed to determine the proportions of therapeutic inertia when treatment targets not achieved in adults T2D at three public health clinics in Malaysia.

**Methods:** The index prescriptions were those when the annual blood tests were reviewed. Prescriptions were verified and classified as 1) no change, 2) stepping up and 3) stepping down. Multivariable logistic regression and sensitive analyses were conducted.

**Results:** At follow-up, 552 participants were available for the assessment of therapeutic inertia (78.9% response rate). The mean (SD) age and diabetes duration were 60.0 (9.9) years and 5.0 (6.0) years, respectively. High therapeutic inertia were observed in oral anti-diabetic (61-72%), anti-hypertensive (34-65%) and lipid-lowering therapies (56-77%), and lesser in insulin (34-52%). Insulin therapeutic inertia was more likely among those with shorter diabetes duration (adjusted OR 0.9, 95% CI 0.87, 0.98). Those who did not achieve treatment targets were less likely to experience therapeutic inertia: HbA1c ≥ 7.0%: adjusted OR 0.10 (0.04, 0.24); BP ≥ 140/90 mmHg: 0.28 (0.16, 0.50); LDL-cholesterol ≥ 2.6 mmol/L: 0.37 (0.22, 0.64).

**Conclusions:** Although therapeutic intensifications were more likely in the present of non-achieved treatment targets but the proportions of therapeutic inertia were high.

**Trial registration:** NCT02730754 https://clinicaltrials.gov/ct2/show/NCT02730754

**Highlights:**

- Probably the first study in Malaysia or Asian countries
- Assessing three main therapeutic inertia among T2D at primary diabetes care
- Proportions of therapeutic inertia were high
- Therapeutic inertia was unlikely in non-achieved treatment targets
- Possible causes of therapeutic inertia require identification and rectification

## 1 Introduction

Managing and achieving optimal control in glycaemia (HbA1c), blood pressure and low-density lipoprotein cholesterol for people with type 2 diabetes (T2D) has been very difficult [1, 2]. Proportions of achieved treatment targets in the world for HbA1c < 7.0% (< 53 mmol/mol) were about 50%, blood pressure < 140/90 mmHg 80% and low-density lipoprotein cholesterol (LDL-cholesterol) 60% [3-6], and it is worse in lower income countries [7-10] and better in a higher income country [11]. In developing countries in Asia, Latin America and Eastern Europe, only 3.6% of T2D patients were able to attain all three recommended targets (blood pressure < 130/80 mmHg, LDL-cholesterol < 100 mg/dl, and HbA1c < 7%)[10]. The same rate was reported as 22% in two polyclinics in Singapore [12]. Without due clinical agility and healthcare system management for T2D, achieving and maintaining optimal treatment targets will face an uphill task and untoward consequences to all [13-16].

Clinical inertia is defined as a failure to initiate or intensify necessary treatments at a timely manner when faced with objective evidence of uncontrolled diseases in the present of clear clinical practice guidelines [17-19]. Therapeutic inertia, although used interchangeably with clinical inertia, focuses more on the prescribed therapies and pharmacological agents, and usually construed as the providers’ failure to increase therapy when treatment targets are not met [20]. Therapeutic inertia could be due to patient-related factors [21], physician-related and healthcare system-related barriers [17]. A combination of these factors may exist and compound effective and efficient diabetes care delivery and clinical consultation between the doctors and patients, patients and facility, and doctors and facility [18, 19]. Some of the principal causes of clinical and therapeutic inertia are doctors’ preference for status quo to avoid uncertainty and risk [22], and impaired communication between doctors and patients [19]. Others include a lack of infrastructure and facility for proper disease monitoring to achieve treatment goals, the mindset of ‘waiting until next visit’ in response to soft rationalizations by patients to avoid treatment intensification, overestimation of care provided, a lack of education and training of the doctors, and practice organization aimed at achieving treatment targets [23, 24].

The problems of therapeutic inertia in diabetes care or a delayed response to poor glycaemic control were about 30-40% or six months to 2 years, respectively [18]. In a recent systematic review, the median time to treatment intensification after at least one HbA1c measurement above target ranged from 0.3 to > 7.0 years [25]. The therapeutic inertia increased with the number of drugs and decreased with increasing HbA1c levels [25]. The UK Clinical Practice Research Datalink (CPRD) reported the median time of basal insulin intensification from initiation was > 4 years, and less than one-third of the eligible T2D [HbA1c ≥ 7.5% (58 mmol/mol)] had their treatment intensified (median time: 3.7 years) [26]. The corresponding therapeutic inertia for hypertension (≥130/80 mmHg) and dyslipidaemia (LDL-C ≥ 2.6 mmol/L or 100 mg/dl) were 46% and 40%, respectively [10].

The evidence on therapeutic inertia has been lacking at primary care setting, especially in low- and middle-income countries and in Asia [25]. Accordingly, this study aimed to determine the proportions and associated factors of therapeutic inertia of anti-diabetic, anti-hypertensive and lipid-lowering therapies when treatment targets not achieved in adults with T2D at public health clinics in Malaysia.

## 2 Methods

This was a cohort study that included baseline and follow-up data from a previous study in 2013 [27], together with the follow-up data from 2014 to 2016. During this period of follow-up, patients received standard diabetes care and clinical services at the respective health clinics. The study was approved by the Medical Research Ethics Committee (MREC), Ministry of Health Malaysia.

### 2.1 Setting and participants

At baseline, participants were sampled consecutively as they came to the clinics over a period of six months. Inclusion criteria at baseline: age 30 years or older, a diagnosis of T2D more than one year ago, and with at least three clinic visits in the previous year. The baseline exclusion criteria were pregnancy or lactating, psychiatric/psychological disorders that could impair judgment and memory, and participants who could not read or understand English, Malay or Mandarin [27]. The participating health clinics were chosen because they serve different sections of the local population. One health clinic (SK) is urban and is visited mainly by patients of Chinese descent, the second is a rural clinic (DK) visited by proportionally more patients of Indian descent than found in a usual public health clinic, and the third clinic (SL) is in a rural and predominantly ethnic Malay residential area. Before the questionnaires were filled in, all participants gave written consent in their preferred language while waiting for a medical consultation with the clinic’s doctor. Trained research assistants interviewed participants at baseline who were not able to self-administer the questionnaires.

### 2.2 Data collection

Baseline demographic data included age, gender, ethnicity, religion, educational level, employment status, monthly income and life event within the past six months [28]. Structured case record forms were used for data collection from the medical records. These included duration of diabetes, HbA1c, diabetes-related complications, blood pressure, lipids, number and type of medication use [27]. At follow-ups, participants were identified by an orange label on their follow-up cards and on their medical records. Follow-up data were retrieved from the medical records on glycaemic control (HbA1c), systolic (SBP) and diastolic blood pressure (DBP), LDL-cholesterol and prescribed medications [27]. These medications included oral anti-diabetic agents (ADA), insulin, oral anti-hypertensive agents (AHA) and lipid-lowering agents (LLA). Non-participants were those who lost to follow-up at the participating clinics and non-attendance for at least more than one year between 2014 and 2016. There was no other injectable ADA besides insulin at the three participating clinics during the period of the study. Every result of HbA1c and LDL-cholesterol, and every reading of SBP and DBP were retrieved. The recommended treatment targets are HbA1c < 7.0% (53 mmol/mol), blood pressure < 140/90 mmHg and LDL-cholesterol < 2.6 mmol/L, respectively [29]. Medications use was retrieved in terms of their names, dose and frequency. The medications prescribed when the annual blood tests were reviewed during the follow-up visit were considered as the index prescription. In the event of absent of an annual blood test, the medications prescribed in the last follow-up visit of the year were considered as the index prescription.

### 2.3 Data analysis

Data analyses were carried out using SPSS software version 25.0 (IBM Corp., Armonk, NY). Comparisons of mean levels were performed using the Student’s t-test or Mann-Whitney U test according to the data distribution, and the Chi-square test was used for proportionate samples between groups. Characteristics of the patients who have and did not have the required data for this study are presented as mean (SD) or median (IQR) for continuous variables, and counts and percentages for nominal variables using descriptive statistics according to the three health clinics.

Assessment of clinical inertia was done for year 2015 because the required medication use data were captured from year 2014, and not possible for year 2016 as the study ended in the third quarter. Index of medication prescription and use of the year were compared to the previous year index prescription. Changes of medication use were classified as 1) no change, 2) stepping up: dose increment or/and replacement with a stronger medication, and 3) stepping down: dose reduction or/and replacement with a weaker medication. All the classification was verified by the author himself and the resident family physicians in SK and SL health clinics. Therapeutic inertia is defined as no change in the medication use in the present of not reaching the recommended treatment targets. Therapeutic inertia was assessed for HbA1c (no change in ADA and insulin uses separately) and LDL-cholesterol (no change in LLA use) by looking at the treatment target in the same year because medication use and change was captured after the tests were reviewed. For those who did not have results for HbA1c and LDL-cholesterol in the same year, the results in the previous year were used. This reflects the actual clinical practice at these health clinics. The therapeutic inertia assessment for blood pressure (no change in AHA use) was assessed by looking for occurrence of persistent SBP/DBP above treatment targets over two occasions with no change of AHA use in the same year. Sensitive analysis was conducted to examine the states of therapeutic inertia for AHA use in the first half and the second half of the year when at least four SBP/DBP measurements were available, and at least two consecutive SBP/DBP measurements that were above the recommended treatment targets in the respective 6-month periods. The results were essentially similar.

Possible clinic and patient’s factors on the therapeutic inertia were further evaluated using the multivariable logistic regression analysis. Besides the clinic, covariates with a *P* value < 0.20 from the univariable analyses were included in the final multivariable analyses. Category ‘no change’ represents therapeutic inertia and category ‘stepping up and stepping down’ was used as the reference group. The final models were re-run with ‘stepping up’ as the reference group and the results did not change the interpretations. We reported the results with the treatment targets entered as one of the covariate, and the reference category of ‘stepping up and stepping down’ as this could account for all reasons of therapeutic changes more clinically meaningful models [30]. Statistical significance was set at *P* < 0.05.

## 3 Results

### 3.1 Cohort characteristics

In total, 552 participants have the required follow-up data for the assessment of therapeutic inertia, representing 78.9% of the baseline sample (n=700). There were no differences in the prescribed medications between the non-participants and participants except LLA use was more among the participants (72% vs. 83%) (Table 1). Compared to the participants, the non-participants were more often treated at the rural Salak Health Clinic, had higher systolic blood pressure, and had dyslipidaemia (Table 1).

**Table 1:**
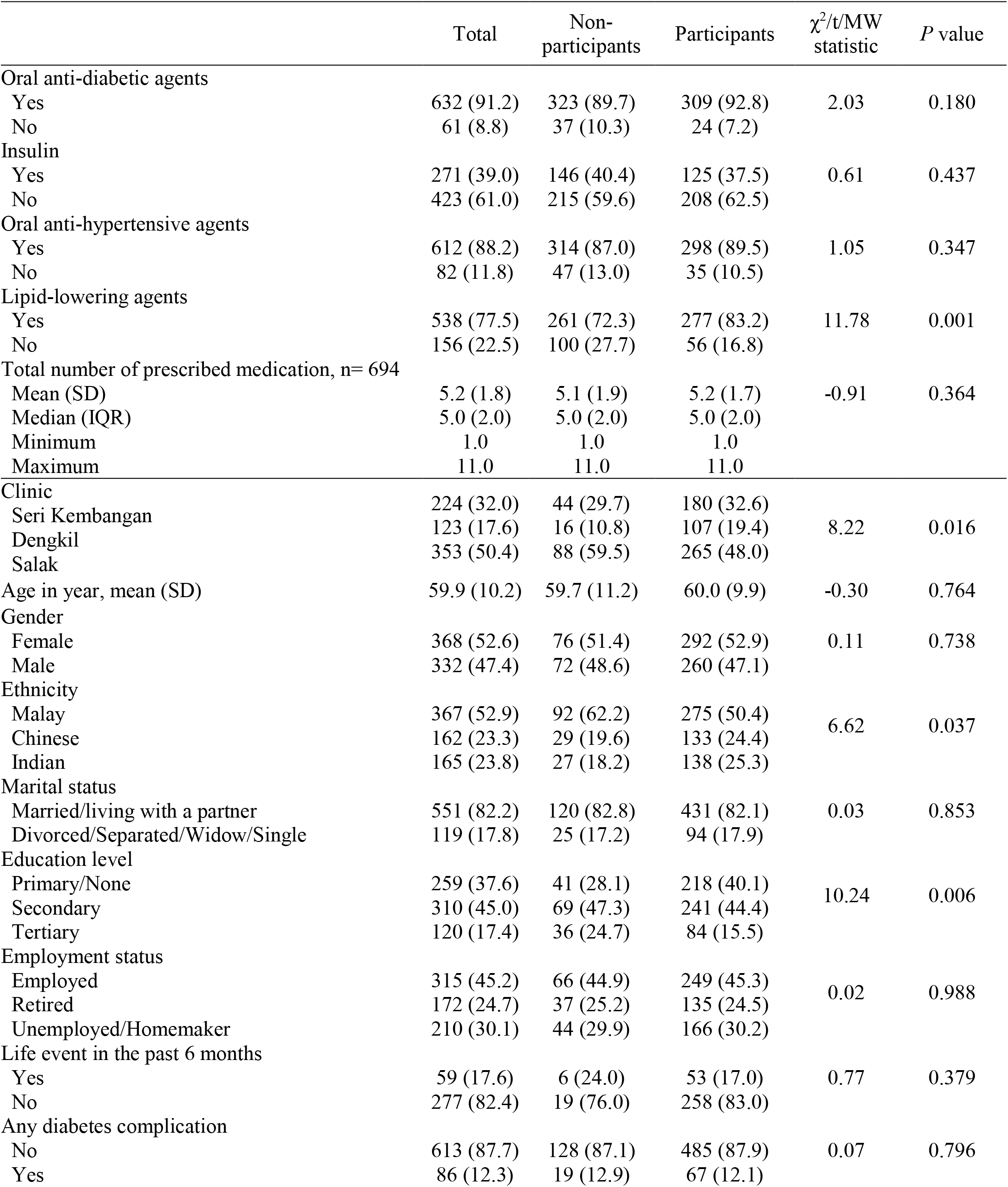
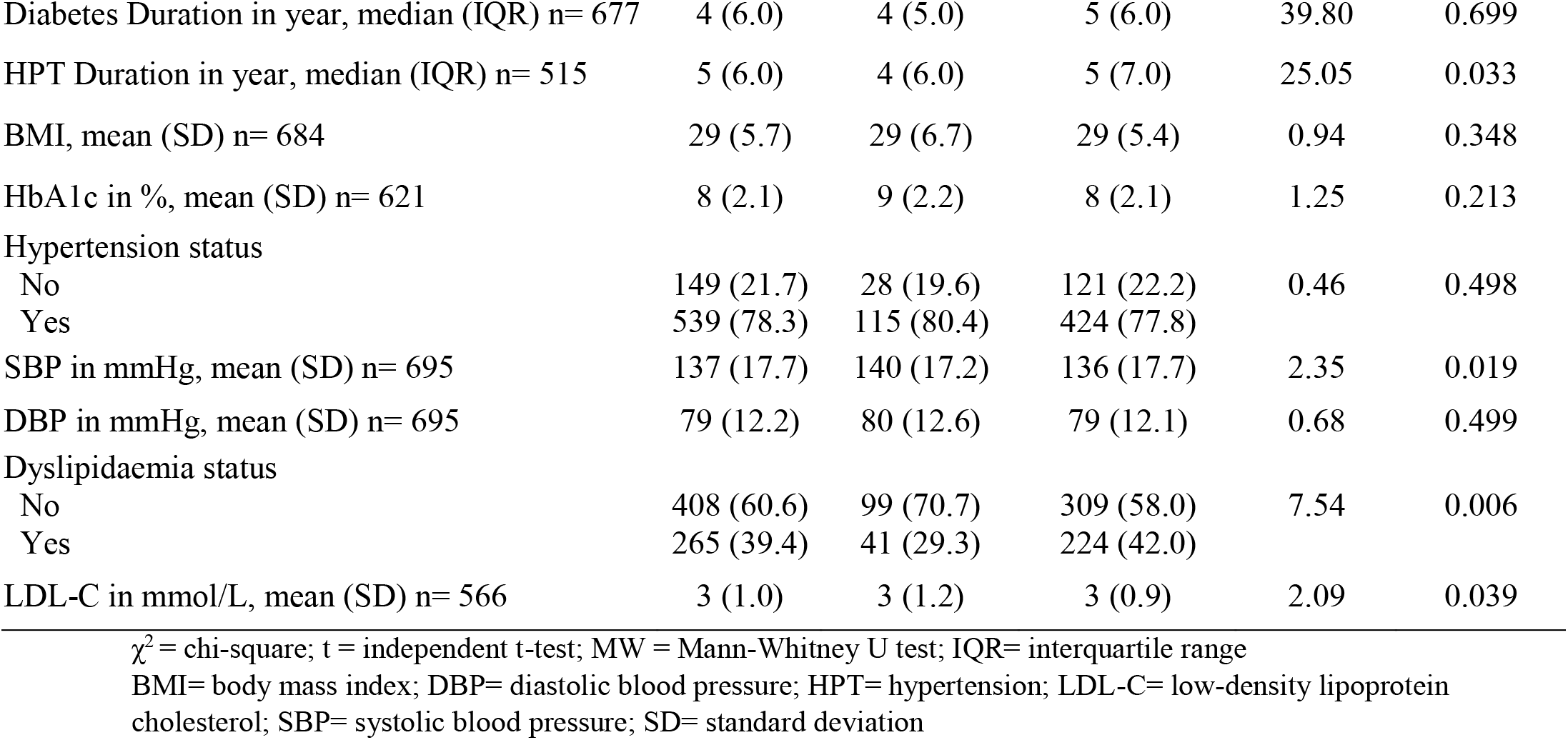
Baseline characteristics between non-participants and participants, n (column %) and total n = 700, unless stated otherwise

### 3.2 Therapeutic inertia in the three clinics

Except for ADA and LLA, there were significant differences in insulin and AHA therapeutic patterns when treatment targets were not achieved at the three health clinics (Figure 1). DK was less likely to have insulin therapy inertia and SK less likely to have AHA inertia compared to the other health clinics. All clinics had high therapeutic inertia in ADA (61-72%) and LLA therapies (56-77%).

**Figure 1:**
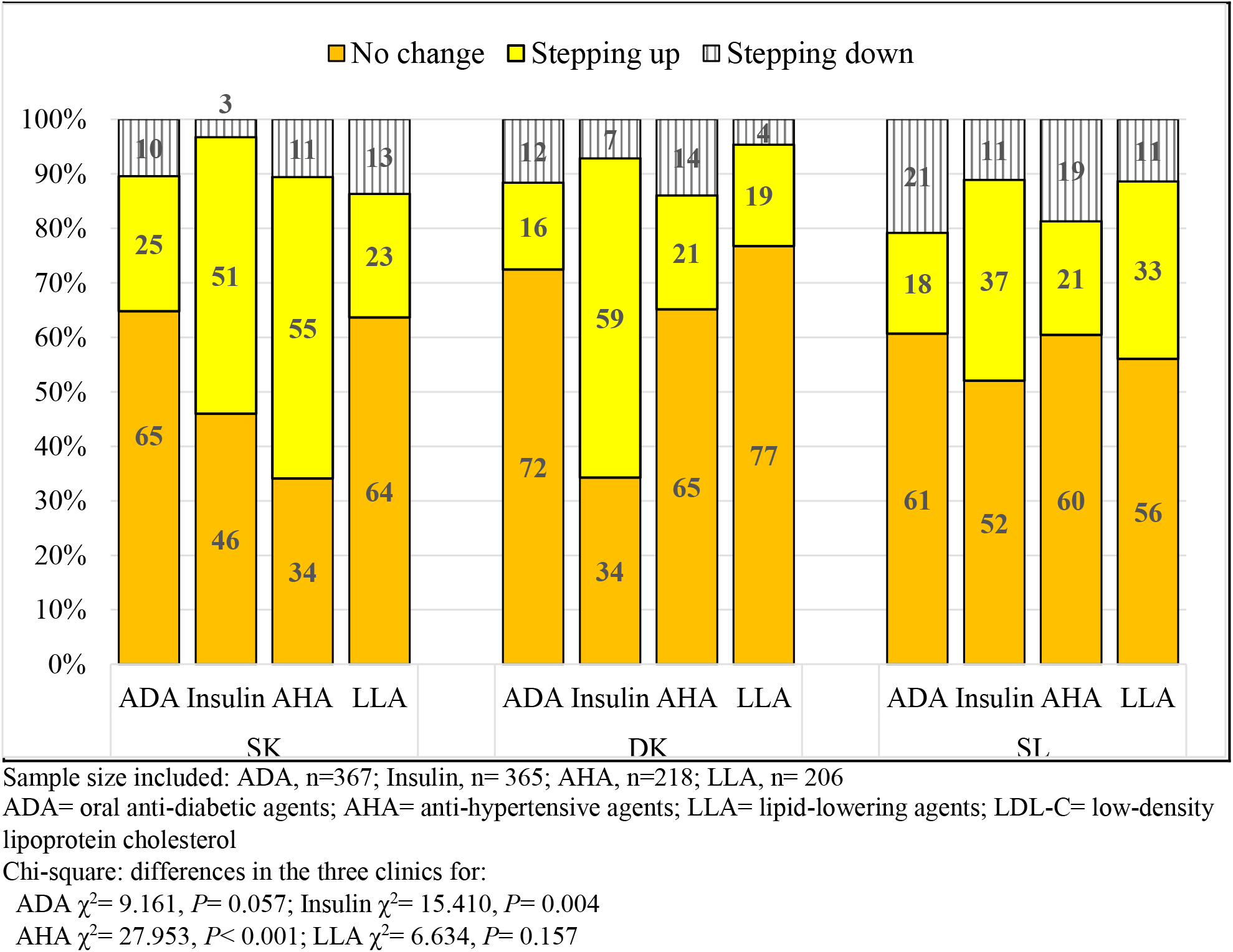
Anti-diabetic, anti-hypertensive and lipid-lowering therapeutic changes when their respective treatment targets were not achieved in year 2015

### 3.3 Anti-diabetics therapy inertia

Both ADA and insulin therapeutic inertia were observed, and ADA therapeutic inertia was worse than the insulin (64% versus 47%) (Table A.1) presumably because of limited dosing range and choices of the oral agents. Patients who were prescribed less medication, had shorter diabetes duration and not having dyslipidaemia were likely to experience ADA therapy inertia (Table A.1). There was no relation between ADA changes and glycaemic control (OR 1.18, 95% CI 0.77, 1.80; *P*= 0.454). Insulin therapeutic inertia was more likely among those with shorter diabetes duration (adjusted OR 0.9, 95% CI 0.87, 0.98) (Table 2).

**Table 2:** Factors associated with insulin therapeutic inertia

| Parameter | Crude Odd Ratio<br>(95% CI) | P value | Adjusted Odd Ratio<br>(95% CI) | P value |
| --- | --- | --- | --- | --- |
| **Insulin therapeutic inertia, n= 284** |  |  |  |  |
| HbA1c < 7.0% * |  |  |  |  |
| HbA1c ≥ 7.0% | 0.09 (0.05, 0.17) | < 0.001 | 0.10 (0.04, 0.24) | < 0.001 |
| Age in year | 1.02 (1.00, 1.04) | 0.039 | 1.03 (0.99, 1.06) | 0.078 |
| Life event in the past 6 months (No*) |  |  |  |  |
| Yes | 1.59 (0.84, 3.01) | 0.159 | 2.01 (0.94, 4.28) | 0.072 |
| Diabetes complications (No*) |  |  |  |  |
| Diabetes complications (Yes) | 0.64 (0.38, 1.07) | 0.086 | 0.75 (0.33, 1.72) | 0.499 |
| Hypertension status (No*) |  |  |  |  |
| Yes | 1.42 (0.94, 2.15) | 0.092 | 1.20 (0.61, 2.35) | 0.595 |
| Diabetes Duration in year | 0.93 (0.90, 0.96) | < 0.001 | 0.92 (0.87, 0.98) | 0.005 |

HbA1c= glycated haemoglobin; CI= confidence interval
\*Referent group
Nagelkerke R Square for this model is 0.29.

### 3.4 Anti-hypertensive therapy inertia

AHA inertia was observed in about half (51%) of the participants with BP > 140/90 mmHg, and only 34% had their AHA therapy stepped up. This inertia happened more among the younger age group, Indians, in active employment, shorter hypertension duration and not diagnosed of hypertension (Table A.2). Those who had at least two consecutive BP > 140/90 mmHg were more likely to experience AHA therapy intensification (Table 3).

**Table 3:** Factors associated with anti-hypertensive therapeutic inertia

| Parameter | Crude Odd Ratio<br>(95% CI) | P value | Adjusted Odd Ratio<br>(95% CI) | P value |
| --- | --- | --- | --- | --- |
| **Anti-hypertensive therapeutic inertia, n= 281** |  |  |  |  |
| At least 2 consecutive BP ≥ 140/90 mmHg in the whole 2015 (No*) | | | | |
| Yes | 0.33 (0.23, 0.47) | < 0.001 | 0.28 (0.16, 0.50) | < 0.001 |
| Health clinic (Seri Kembangan*) |  |  |  |  |
| Dengkil | 4.09 (2.29, 7.31) | < 0.001 | 4.79 (2.03, 11.27) | < 0.001 |
| Salak | 1.75 (1.19, 2.58) | 0.004 | 1.55 (0.71, 1.00) | 0.272 |
| Age in year | 0.96 (0.94, 0.98) | < 0.001 | 0.97 (0.93, 1.00) | 0.054 |
| Ethnicity (Malay*) |  |  |  |  |
| Chinese | 0.61 (0.40, 0.93) | 0.022 | 0.63 (0.30, 1.32) | 0.222 |
| Indian | 1.38 (0.88, 2.18) | 0.163 | 1.03 (0.46, 2.28) | 0.946 |
| Employment status (Employed*) |  |  |  |  |
| Unemployed | 0.70 (0.46, 1.08) | 0.104 | 1.48 (0.72, 3.06) | 0.287 |
| Retired/house wife | 0.49 (0.32, 0.76) | 0.001 | 1.03 (0.48, 2.22) | 0.944 |
| Life event in the past 6 months (No*) |  |  |  |  |
| Yes | 0.64 (0.35, 1.18) | 0.151 | 1.01 (0.47, 2.16) | 0.988 |
| Diabetes complications (No*) |  |  |  |  |
| Diabetes complications (Yes) | 0.69 (0.41, 1.16) | 0.162 | 1.05 (0.40, 2.80) | 0.917 |
| Hypertension status (No*) |  |  |  |  |
| Yes | 0.48 (0.30, 0.77) | 0.003 | 0.56 (0.22, 1.40) | 0.214 |
| Diabetes Duration in year | 0.95 (0.92, 0.98) | 0.002 | 1.01 (0.95, 1.06) | 0.860 |
| Hypertension Duration in year | 0.96 (0.93, 0.99) | 0.007 | 0.99 (0.94, 1.05) | 0.750 |

BP= blood pressure; CI= confidence interval
\*Referent group
Nagelkerke R Square for this model is 0.28.

### 3.5 Lipid lowering therapy inertia

Therapeutic inertia in LLA was observed in 61% of those who had uncontrolled LDL-cholesterol, and this was more common among those having hypertension on top of T2D (Table A.3). However, no factor was an independent risk factor to LLA therapy inertia after adjusting for LDL-cholesterol treatment target (Table 4). Those who had not achieved LDL-cholesterol treatment target < 2.6 mmol/L were more likely to experience lipid-lowering therapy intensification.

**Table 4:** Factors associated with lipid lowering therapeutic inertia

| Parameter | Crude Odd Ratio<br>(95% CI) | *P* value | Adjusted Odd Ratio<br>(95% CI) | *P* value |
| --- | --- | --- | --- | --- |
| **Lipid lowering therapeutic inertia, n= 332** |  |  |  |  |
| LDL-cholesterol < 2.6 mmol/L * |  |  |  |  |
| LDL-cholesterol ≥ 2.6 mmol/L | 0.35 (0.21, 0.57) | < 0.001 | 0.37 (0.22, 0.64) | < 0.001 |
| Health clinic (Seri Kembangan*) |  |  |  |  |
| Dengkil | 1.31 (0.73, 2.34) | 0.365 | 1.11 (0.42, 2.89) | 0.836 |
| Salak | 0.61 (0.40, 0.93) | 0.021 | 0.74 (0.32, 1.74) | 0.490 |
| Age in year | 1.02 (1.00, 1.04) | 0.042 | 1.02 (0.99, 1.05) | 0.303 |
| Education level (Primary/None*) |  |  |  |  |
| Secondary | 0.70 (0.47, 1.06) | 0.091 | 0.88 (0.48, 1.62) | 0.690 |
| Tertiary | 0.72 (0.51, 1.60) | 0.720 | 1.34 (0.60, 3.01) | 0.481 |
| Employment status (Employed*) |  |  |  |  |
| Unemployed | 1.04 (0.67, 1.59) | 0.875 | 1.22 (0.59, 2.53) | 0.595 |
| Retired/house wife | 1.59 (0.98, 2.59) | 0.062 | 1.11 (0.61, 2.04) | 0.727 |
| Marital status |  |  |  |  |
| Married/living with a partner* |  |  |  |  |
| Divorced/Separated/Widow/Single | 1.58 (0.92, 2.69) | 0.095 | 1.41 (0.67, 2.95) | 0.366 |
| Hypertension status (No*) |  |  |  |  |
| Yes | 1.40 (0.90, 2.17) | 0.132 | 1.45 (0.83, 2.55) | 0.192 |

LDL-cholesterol = low density lipoprotein-cholesterol; CI= confidence interval
\*Referent group
Nagelkerke R Square for this model is 0.12.

## 4 Discussion

This study examined the therapeutic patterns of ADA, AHA and LLA in adults with T2D after three years of regular primary diabetes care at three public health clinics in Malaysia. The findings show some similarity and differences in the therapeutic changes of these medications in the three health clinics when treatment targets were not achieved. Although multivariable analyses show that treatment intensifications were more likely in the face of uncontrolled diseases, but the proportions of therapeutic inertia were high in hyperglycaemia (up to 64%), hypertension (51%) and high LDL-cholesterol (61%). These proportions were still considered high after taking into account the possible contraindications, intolerance and refusals to medications of about 15 to 20% based on the authors’ experience and early studies [31, 32].

High prevalent of therapeutic inertia in ADA in all three clinics might indicate limitation in the dosing of the oral medications when maximum doses have been prescribed and with the restricted choices of ADA [33]. The commonly available ADA are metformin and gliclazide, with additional one to two drugs from the newer classes of dipeptidyl peptidase-4 (DPP-4) inhibitors, which are restricted to the resident family physicians use. Therefore, the observed therapeutic inertia could be a result of clinical inertia in timely referral for other ADA or insulin initiation. It was also noted that patients who were relatively healthier (shorter diabetes duration, no comorbid such as dyslipidaemia and fewer number of medication) were more likely to experience ADA inertia [17]. In contrast to dose increment limitation in ADA, insulin has wider dosing possibility and this had resulted in relatively lower insulin inertia, and recorded the highest stepping-up rate among all the studied therapies in this study. The observed rate of insulin intensification was similar to the Canadian specialist treating non-insulin-requiring patients in year 2000 [34]. Patients with shorter diabetes duration of T2D were more likely to experience insulin inertia. This might be construed that both the patients and doctors preferred working harder on non-pharmacological means or ADA before insulin dose increment.

AHA therapeutic patterns noted a wide variations across the three clinics, and different clinic was one of the significant factors that showed an independent effect on AHA inertia in the multivariable model. DK was most likely to have AHA inertia compared to the other clinics. These could be due to the differences in healthcare system-related factors such as having a dedicated team and consultation room at SK [33]. This might contribute to a more effective consultation with patients that improved doctor-patient communication. Other possible causes include having competent knowledge in hypertension, and familiarity with newer AHA that are beneficial in T2D. Therapeutic inertia in AHA could also be due to the patient-related factors such as denial and refusal of treatment intensification due to non-experiencing of symptoms of hypertension or disease progression [17]. Although this study did not examine the time to anti-hypertensive treatment intensification since two consecutive BP > 140/90 mmHg had been recorded, it was likely that at least 40% experienced AHA inertia for longer than one year based on the sensitive analysis conducted.

Multivariable analysis did not reveal any significant contributing factor towards LLA inertia except that of achieving the LDL-cholesterol target. This was an assuring finding but the proportion of therapeutic inertia was highest in LLA among the three therapies. This might indicate that the causes were less of physician-related but more of patient-related or health system-related. The availability of LLA to the clinics’ doctors was generally restricted to three types of LLA namely lovastatin, simvastatin and gemfibrozil, with atorvastatin and fenofibrate were further available with endorsement by the resident family physicians with a specialist status. Patient-related causes might be statin intolerance and concern about statins worsen the glycaemic control [35]. Lessons from controlled clinical trials suggested that a combination of good patient education and support, and clear treatment strategies might reduce clinical inertia [17]. However, this may not ensure timely treatment intensification by the attending doctors who perceive time constraint in consultation [36] and concern about statin adverse effects.

### 4.1 Strength and limitations

This was probably one of the first study in Malaysia or Asia assessing all three main therapeutic inertia among T2D at primary care in a prospective manner. Good sample size and representative samples of the participants [27] render the study sufficient power to produce the answers. This study was subjected to several potential limitations. This study did not classify diabetes pharmacological regimen as a whole but ADA to insulin changes separately. Thus, escalation of treatment from ADA to insulin therapy in the event of HbA1c ≥ 7.0% was not recorded. Therefore, insulin therapeutic inertia provided an estimate that was closer to the actual clinical practice because from none to insulin initiation was captured. Treatment targets used did not reflect risk profiles and customized target levels for the patients. This may lead to underestimation of the proportions of therapeutic inertia because majority of the adults T2D at the primary care setting were of the lower risk and in early stages of diseases, and treatment intensification should occur at lower target levels [37, 38]. Similarly, underestimation of the proportions of therapeutic inertia is likely in the participants especially in ADA and LLA since the non-participants were prescribed less of these medications at baseline. The three participating health clinics in this study are of the medium scale with resident family physicians and doctors, and situated in a developed state of Selangor across urban, sub-rural and rural regions. This limits generalizability of the results to other clinics of different scales, without resident doctors or in the more remote areas in the country that may have different healthcare systems and delivery at the meso- and micro-levels.

## 5 Conclusion

Although therapeutic intensifications were more likely in the present of non-achieved glycaemic, blood pressure and LDL-cholesterol treatment targets but the proportions of adults with T2D who faced therapeutic inertia were high at primary diabetes care in Malaysia. Possible causes of therapeutic inertia require urgent identification and rectification to improve disease control [39-41]. Delay in treatment intensification for T2D and prolonged suboptimal control of hypertension and high LDL-cholesterol in people with T2D will lead to detrimental health implications and costly consequences.

## Data Availability

Data will be made available upon request.

## Conflict of interest

The authors declare that this research was conducted in the absence of any commercial or financial relationships that could be construed as a potential conflict of interest.

## Acknowledgements

We would like to thank the Director General of Health Malaysia for his permission to publish this article. We thank Universiti Putra Malaysia (UPM) and the Ministry of Higher Education, Malaysia for their support in sponsoring the Ph.D. study and living allowances for Boon-How Chew. We acknowledge Prof. Dr. Guy E.H.M. Rutten from the University of Utrecht, Julius Center for Health Sciences and Primary Care and Ass. Prof. Dr. Rimke C. Vos from Leiden University Medical Center for participating in the larger EDDMQOL-Pro study. We would like to thank Associate Professor Dr. Sazlina Sharif Ghazali, previous Head of the Department of Family Medicine UPM, Dr. Aaron Fernandez of the Psychiatry Department UPM, Dr. Dr. Noor-Hasliza Hassan at Dengkil Health Clinic, and the Petaling and Sepang District Health Offices for their permission and assistance in this study.

## Funding

This work was partly supported by the Ministry of Higher Education Malaysia (grant numbers KPT(BS)730928075371, 2013) and Universiti Putra Malaysia’s journal publication awards (award numbers UPM/TNCPI/RMC/5.1.17/F5(16)). The funding bodies had no role in the design of the study or in the collection, analysis, or interpretation of data or in writing the manuscript.

## Notes

### Competing Interest Statement

The authors have declared no competing interest.

### Clinical Trial

NCT02730754

### Author Declarations

All relevant ethical guidelines have been followed and any necessary IRB and/or ethics committee approvals have been obtained.

Any clinical trials involved have been registered with an ICMJE-approved registry such as ClinicalTrials.gov and the trial ID is included in the manuscript.

